# A cohort of patients with COVID-19 in a major teaching hospital in Europe

**DOI:** 10.1101/2020.04.29.20080853

**Authors:** Alberto M. Borobia, Antonio J. Carcas, Francisco Arnalich, Rodolfo Álvarez-Sala, Jaime Montserrat, Manuel Quintana, Juan Carlos Figueira, Rosario M. Torres Santos-Olmo, Julio García-Rodríguez, Alberto Martín-Vega, Antonio Buño, Elena Ramírez, Gonzalo Martínez-Alés, Nicolás García-Arenzana Les, M. Concepción Núñez, Milagros Martí-de-Gracia, Francisco Moreno Ramos, Francisco Reinoso-Barbero, Alejandro Martin-Quiros, Angélica Rivera Núñez, Jesús Mingorance, Carlos J Carpio Segura, Daniel Prieto Arribas, Esther Rey Cuevas, Concepción Prados Sánchez, Juan J. Rios, Miguel A. Hernán, Jesús Frías, José R Arribas, for the COVID@HULP Working Group

**Author notes:** **Corresponding authors**: Alberto M. Borobia. Clinical Pharmacology Department, Antonio J. Carcas. Clinical Pharmacology Department, José R. Arribas. Internal Medicine Department. La Paz University Hospital, Paseo de la Castellana, 261, 28046-Madrid (SPAIN). A complete list of the members of the COVID@HULP Working Group is provided in the Supplementary Appendix.

## Abstract

**BACKGROUND:** Since the confirmation of the first patient infected with SARS-CoV-2 in Spain in January 2020, the epidemic has grown rapidly, with the greatest impact on the Madrid region. This article describes the first 2226 consecutive adult patients with COVID-19 admitted to the La Paz University Hospital in Madrid.

**METHODS:** Our cohort included all consecutively admitted patients who were hospitalized and who had a final outcome (death or discharge) in a 1286-bed hospital of Madrid (Spain) from February 25^th^ (first case admitted) to April 19^th^, 2020. Data was entered manually into an electronic case report form, which was monitored prior to the analysis.

**RESULTS:** We consecutively included 2226 adult patients admitted to the hospital who either died (460) or were discharged (1766). The patients’ median age was 61 years; 51.8% were women. The most common comorbidity was arterial hypertension (41.3%). The most common symptom on admission were fever (71.2%). The median time from disease onset to hospital admission was 6 days. Overall mortality was 20.7% and was higher in men (26.6% vs 15.1%). Seventy-five patients with a final outcome were transferred to the ICU (3.4%). Most patients admitted to the ICU were men, and the median age was 64 years. Baseline laboratory values on admission were consistent with an impaired immune-inflammatory profile.

**CONCLUSIONS:** We provide a description of the first large cohort of hospitalized patients with COVID-19 in Europe. Advanced age, male gender, the presence of comorbidities and abnormal laboratory values were more common among the patients with fatal outcomes.

## INTRODUCTION

After the United States, Spain has the second highest number of confirmed infections with severe acute respiratory syndrome coronavirus 2 (SARS-CoV-2) worldwide. The first patient in Spain was confirmed on January 31st, 2020^1^ and in the Madrid region on February 25th, 2020, an area 6.7 million individuals^2^ where the epidemic had the greatest impact. The number of confirmed cases in Madrid was 58,819 as of April 25 (26.3% of cases in Spain)^3^, with a maximum of 3419 new cases on March 30.

The progression of the outbreak in Madrid is quite similar to that observed in the most affected areas in western countries such as the Lombardia region in Italy, or New York city in US. The healthcare systems of these areas are under very high stress and the mortality rates (per 100,000 inhabitants) are high: 132 in Lombardia, 140 in New York City and 190 in Madrid (as of 25^th^ of April).^4-6^

La Paz University Hospital is a major teaching hospital that serves a catchment area of 527,366 individuals in the north of Madrid. Shortly after the outbreak, hospital’s operations were adapted to cope with the surge in COVID-19 cases. By April 25, the hospital had admitted over 2500 patients with COVID-19, one of the largest single-site cohorts in Europe. The clinical information of these patients was collected using a standardized protocol.

Here, we describe the first consecutive 2226 adult patients with a confirmed SARS-CoV-2 infection in La Paz University Hospital and who had been discharged or died by April 19.

## METHODS

### Study population

Our study includes all individuals 18 years or older who were hospitalized in the wards (or emergency department, due to lack of available beds in the wards) of La Paz University Hospital with a COVID-19 diagnosis and who had been discharged or dead by April 19^th^. Patients discharged from the emergency department after less than a 24-hour stay were not considered hospitalized and were not included in this analysis.

### Data collection

A modified version of the electronic case record form (eCRF) for severe acute respiratory infections by the World Health Organization/International Severe Acute Respiratory and Emerging Infection Consortium was used^7^. Our eCRF includes 372 variables grouped into demography, medical history, infection-exposure history, symptoms, complications, treatments (excluding clinical trials) and evolution of the disease during hospitalization (Supplementary material).

Clinical data were collected by direct extraction when this was possible, or by manual and individual review of the patients’ electronic clinical records, including clinical notes (DXC-HCIS-Healthcare Information System). Clinical data at hospital admission included age, sex, smoking status, transmission, comorbidities, symptoms on admission, respiratory status and time from disease onset. Complications during hospitalization and admission to ICU were also recorded. This data collection effort has been carried out by a volunteer team of resident doctors and senior medical students. Data monitoring was conducted by the Central Clinical Research Unit of our hospital.

Laboratory results (hematology, biochemistry, microbiology) were extracted from different hospital data management systems. Medications during hospitalization were extracted from the electronic prescription system (EPSs).

### Statistical analysis

Continuous variables are presented as mean and standard deviation (SD) or median and interquartile range (IQR). Categorical variables are listed as numbers and percentages (%), respectively. We performed the statistical calculations using R (version 3.4.0)^8^.

The study was approved by the Research Ethics Committee of La Paz University Hospital (PI-4072) and by the Spanish Agency of Medicines and Medical Devices (HUL-AIN-2020-01) and was registered in the European Union Electronic Register of Post-Authorization Studies (EUPAS34331).

## RESULTS

A total of 3127 patients were consecutively treated in the emergency department of La Paz University Hospital between Feb 25^th^ and April 19^th^, 2020. Of these, 2226 adult patients were hospitalized and either died (460, 20.7%) or were discharged (1766, 79.3%) and therefore included in our analysis. Figure 1 shows occupancy with by COVID-19 patients over time, with a peak of 1033 beds (compared with 1268 pre-COVID beds for the entire hospital) of which 106 were in ICU (compared with 30 pre-COVID ICU beds).

**Figure 1.**
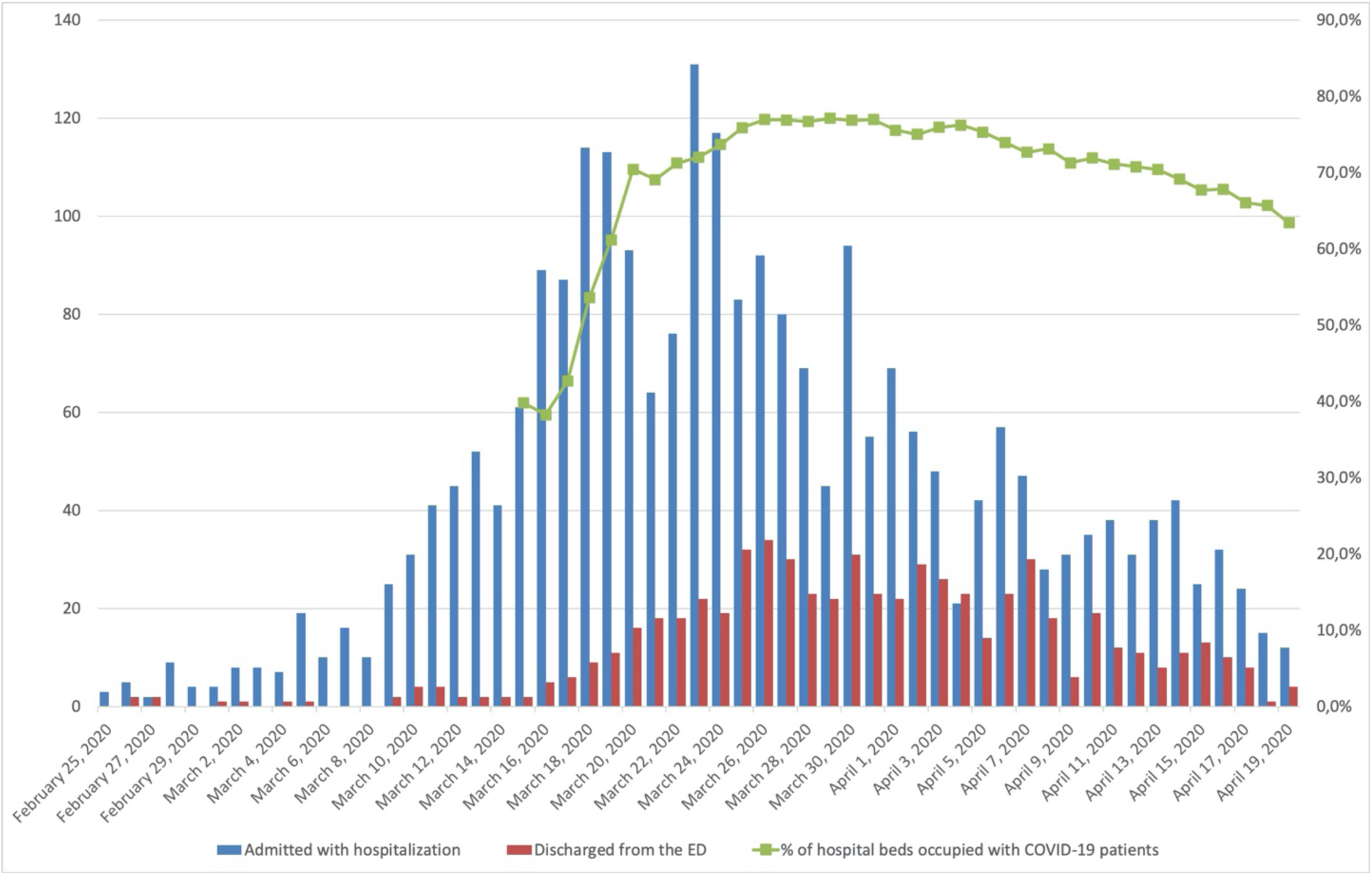
Patients attending emergency department per day between Feb 25th, 2020 and 19th April, 2020. In blue patients admitted with hospitalization and in red patients discharged from the emergency department. Green line represents *%* of hospital beds (including ICU) occupied with COVID-19 patients.

The median time from clinical onset to hospital admission was 6 days (IQR, 3–9). At admission, patients had a median age of 61 (IQR, 46–78) years, 52% were women, 41% had hypertension, 19% had chronic heart disease, and 17% had diabetes mellitus. The most common symptoms on admission were fever, cough and dyspnea. The median oxygen saturation on admission was 95% (IQR, 92–97). The most common complications during the hospitalization were acute confusional syndrome, acute kidney failure and acute respiratory distress syndrome (Table 1).

**Table 1:** Characteristics on Admission, and Complications during Hospitalization

| Variable | Total<br>(n=2226) | Deaths<br>(n=460) | Live discharges<br>(n=1766) |
| --- | --- | --- | --- |
| **Median age, years (IQR)** | 61 (46–78) | 82.5 (76–87) | 56 (42–71) |
| **Sex, n (%)** |  |  |  |
| Male | 1074 (48.2) | 286 (62.2) | 788 (44.6) |
| Female | 1152 (51.8) | 174 (37.8) | 978 (55.4) |
| **Nursing home resident** | 709 (31.9) | 125 (27.2) | 584 (33.1) |
| **Current smokers, n (%)** | 157 (7.1) | 44 (9.6) | 113 (6.4) |
| **Comorbidities (at least one), n (%)** | 1747 (78.5) | 448 (97.4) | 1299 (73.6) |
| Arterial hypertension | 920 (41.3) | 318 (69.1) | 602 (34.1) |
| Chronic heart disease | 429 (19.3) | 195 (42.4) | 234 (13.3) |
| Diabetes mellitus | 381 (17.1) | 157 (34.1) | 224 (12.7) |
| Rheumatological disease | 268 (12.0) | 80 (17.4) | 188 (10.6) |
| Solid malignant disease | 252 (11.3) | 93 (20.2) | 159 (9.0) |
| Obesity | 242 (10.9) | 66 (14.3) | 176 (10.0) |
| Chronic kidney disease | 174 (7.8) | 94 (20.4) | 80 (4.5) |
| Chronic obstructive pulmonary disease | 153 (6.9) | 65 (14.1) | 88 (5.0) |
| Other chronic lung diseases | 143 (6.4) | 49 (10.7) | 94 (5.3) |
| Hematological malignant disease | 133 (6.0) | 46 (10.0) | 87 (4.9) |
| Asthma | 115 (5.2) | 17 (3.7) | 98 (5.5) |
| Liver disease | 89 (4.0) | 23 (5.0) | 66 (3.7) |
| HIV infection | 13 (0.6) | 4 (0.9) | 9 (0.5) |
| **Symptoms on admission, n (%)** |  |  |  |
| Fever | 1585 (71.2) | 325 (70.7) | 1260 (71.3) |
| Cough | 1373 (61.7) | 228 (49.6) | 1145 (64.8) |
| Dyspnea | 1108 (49.8) | 278 (60.4) | 830 (47.0) |
| Myalgia | 596 (26.8) | 51 (11.1) | 545 (30.9) |
| Diarrhea | 484 (21.7) | 62 (13.5) | 422 (23.9) |
| Headache | 427 (19.2) | 24 (5.2) | 403 (22.8) |
| Sputum production | 320 (14.4) | 80 (17.4) | 240 (13.6) |
| Nausea or vomiting | 299 (13.4) | 42 (9.1) | 257 (14.6) |
| Anosmia | 284 (12.8) | 5 (1.1) | 279 (15.8) |
| **Respiratory status** |  |  |  |
| Median SatO<sub>2</sub> on admission. % (IQR) | 95 (92–97) | 90 (85–93.3) | 96 (93–97) |
| SatO<sub>2</sub> <90% on admission, n (%) | 234 (10.5) | 126 (27.4) | 108 (6.1) |
| **Median time from disease onset to hospital admission, days (IQR)** | 6 (3–9) | 4 (2–7) | 7 (3–10) |
| **Complications during hospitalization** |  |  |  |
| Acute confusional syndrome | 197 (8.8) | 149 (32.4) | 48 (2.7) |
| Acute kidney failure | 173 (7.8) | 120 (26.1) | 53 (3.0) |
| Acute respiratory distress syndrome | 109 (4.9) | 94 (20.4) | 15 (0.8) |
| Bacterial pneumonia | 64 (2.9) | 45 (9.8) | 19 (1.1) |
| Acute heart failure | 51 (2.3) | 34 (7.4) | 17 (1.0) |
| Arrhythmia | 46 (2.1) | 33 (7.2) | 12 (0.7) |
| **Treatment regimens, n (%)** |  |  |  |
| Hydroxychloroquine + Azythromycine | 706 (31.7) | 180 (39.1) | 526 (29.8) |
| Hydroxychloroquine | 545 (24.5) | 163 (35.4) | 382 (21.6) |
| Hydroxychloroquine + Lopinavir/ritonavir | 116 (5.2) | 23 (5.0) | 93 (5.3) |
| Hydroxychloroquine + Azythromycine +<br>Lopinavir/ritonavir | 69 (3.1) | 23 (5.0) | 46 (2.6) |
| Lopinavir/ritonavir | 19 (0.8) | 3 (0.6) | 16 (0.9) |

Abbreviations: HIV, human immunodeficiency virus; IQR, interquartile range; SatO<sub>2</sub>, oxygen saturation

The majority of patients received a treatment with presumed antiviral activity against SARS-CoV-2. The most frequent combination was Hydroxychloroquine + Azythromycine followed by Hydroxychloroquine alone

In the moment of the analysis, 237 patients had been admitted in the ICU, 116 remain in the ICU, 55 have died, 20 were discharged from the hospital and 46 remain in an acute hospital bed. Table 2 shows the demographic characteristics, comorbidities and respiratory status on the day of admission to the emergency department of the 75 patients (3.4%) transferred to the intensive care unit (ICU) who had been discharged or died by April 19^th^. Compared with the entire cohort, patients admitted to the ICU were older (median 64 vs 61 years), had a higher male/female ratio (3.2 vs 0.93) and higher prevalence of hypertension (52 vs 41.3%), obesity (30.7% vs 10.9%), diabetes mellitus (28.0% vs 17.1%) and chronic obstructive pulmonary disease (17.3 vs 6.9).

**Table 2:**
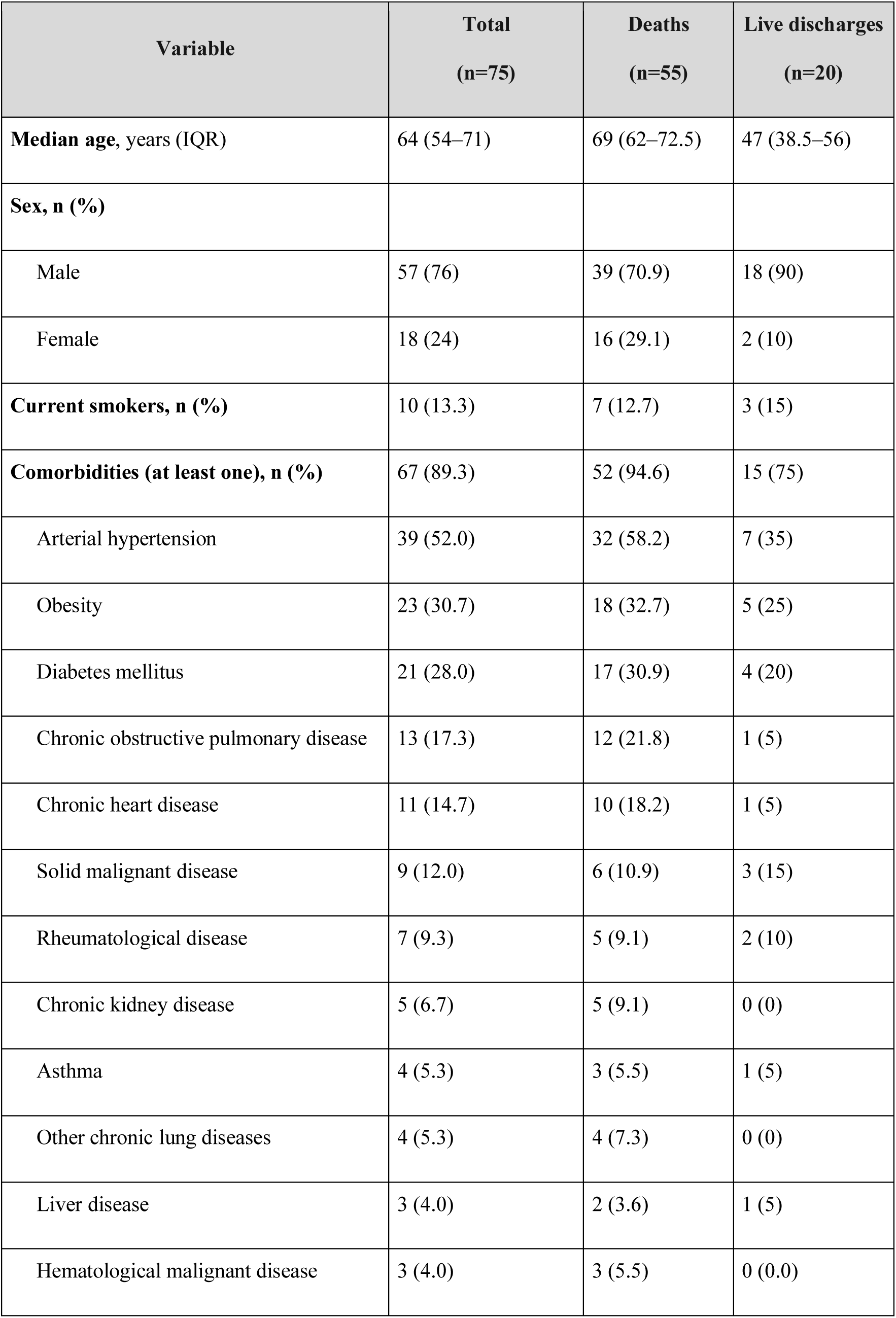
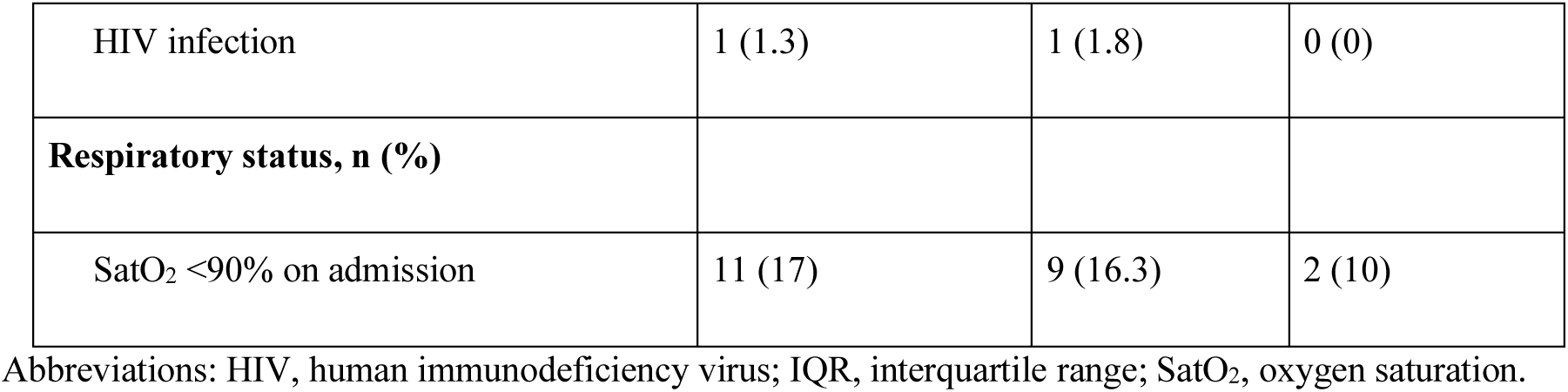
Demographic, Comorbidities and Respiratory Status on Admission of the ICU Patients

| Variable | Total<br>(n=75) | Deaths<br>(n=55) | Live discharges<br>(n=20) |
| --- | --- | --- | --- |
| **Median age, years (IQR)** | 64 (54–71) | 69 (62–72.5) | 47 (38.5–56) |
| **Sex, n (%)** |  |  |  |
| Male | 57 (76) | 39 (70.9) | 18 (90) |
| Female | 18 (24) | 16 (29.1) | 2 (10) |
| **Current smokers, n (%)** | 10 (13.3) | 7 (12.7) | 3 (15) |
| **Comorbidities (at least one), n (%)** | 67 (89.3) | 52 (94.6) | 15 (75) |
| Arterial hypertension | 39 (52.0) | 32 (58.2) | 7 (35) |
| Obesity | 23 (30.7) | 18 (32.7) | 5 (25) |
| Diabetes mellitus | 21 (28.0) | 17 (30.9) | 4 (20) |
| Chronic obstructive pulmonary disease | 13 (17.3) | 12 (21.8) | 1 (5) |
| Chronic heart disease | 11 (14.7) | 10 (18.2) | 1 (5) |
| Solid malignant disease | 9 (12.0) | 6 (10.9) | 3 (15) |
| Rheumatological disease | 7 (9.3) | 5 (9.1) | 2 (10) |
| Chronic kidney disease | 5 (6.7) | 5 (9.1) | 0 (0) |
| Asthma | 4 (5.3) | 3 (5.5) | 1 (5) |
| Other chronic lung diseases | 4 (5.3) | 4 (7.3) | 0 (0) |
| Liver disease | 3 (4.0) | 2 (3.6) | 1 (5) |
| Hematological malignant disease | 3 (4.0) | 3 (5.5) | 0 (0.0) |
| HIV infection | 1 (1.3) | 1 (1.8) | 0 (0) |
| **Respiratory status, n (%)** |  |  |  |
| SatO<sub>2</sub> <90% on admission | 11 (17) | 9 (16.3) | 2 (10) |

Abbreviations: HIV, human immunodeficiency virus; IQR, interquartile range; SatO<sub>2</sub>, oxygen saturation.

Table 3 shows the mortality by age group and sex in the 2226 patients. Overall mortality was 26.6% in males and 15.1% in females. Mortality increased with age, reaching over 60% for patients over 80 years of age.

**Table 3:** Mortality Distribution by Age Group and gender

|  | Male |  | Female |  | Total |  |
| --- | --- | --- | --- | --- | --- | --- |
| Age group | n | Mortality (%) | n | Mortality (%) | n | Mortality (%) |
| 18-29 years | 59 | 0.0 | 100 | 1.0 | 159 | 0.6 |
| 30-39 years | 86 | 0.0 | 133 | 0.0 | 219 | 0 |
| 40-49 years | 120 | 1.7 | 167 | 1.2 | 287 | 1.5 |
| 50-59 years | 166 | 4.8 | 199 | 3.0 | 365 | 3.8 |
| 60-69 year | 120 | 20.0 | 152 | 7.9 | 327 | 11 |
| 70-79 years | 202 | 41.1 | 156 | 25.0 | 358 | 34.1 |
| 80-89 years | 212 | 62.7 | 179 | 41.3 | 391 | 52.9 |
| ≥90 years | 54 | 66.7 | 66 | 60.6 | 120 | 63.3 |

Table 4 shows the laboratory findings at admission for the entire cohort and for the ICU subgroup. In the cohort, baseline creatine kinase, creatinine, D-dimer, ferritin, lactate dehydrogenase, procalcitonin, C-reactive protein and high-sensitivity cardiac troponin I levels and prothrombin times were higher among the non-survivors than the survivors. In the cohort admitted to the ICU, the most notable differences with the whole cohort were higher levels of D-dimer, ferritin, C reactive protein and troponin. Also, lymphocyte count, procalcitonin and C-reactive protein levels at hospital admission were clearly higher in the patients who died in comparison with those surviving within the ICU cohort.

**Table 4:** Laboratory Findings on Admission

| Variable, median (IQR) | n with data | Total | Deaths | Live discharges |
| --- | --- | --- | --- | --- |
| **Entire cohort** |  | **n=2226** | **n=460** | **n=1766** |
| Lymphocyte count, $\times 10^9/L$<br>(NR, 1.1–4.5) | 1805 | 0.93 (0.91–0.97) | 0.68 (0.64–0.74) | 1.02 (0.99–1.06) |
| ALT, U/L<br>(NR, 0–35) | 1526 | 31 (30–33) | 28 (27–31) | 32 (31–34) |
| Albumin g/dL<br>(NR, 2.9–5.2) | 1200 | 4.2 (4.2–4.3) | 3.9 (3.9–4) | 4.3 (4.3–4.4) |
| Creatine kinase, U/L<br>(NR, 40–280) | 1241 | 100 (95–105) | 143 (121–164) | 88 (83–95) |
| Creatinine mg/dL<br>(NR, 0.7–1.3) | 1602 | 0.81 (0.8–0.83) | 1.05 (0.99–1.13) | 0.77 (0.75–0.78) |
| D-dimer, ng/mL<br>(NR, 0–500) | 982 | 720 (680–758) | 1590 (1309–1952) | 608 (560–648) |
| Serum ferritin, ng/L<br>(NR, 22–322) | 599 | 346 (312–393) | 588 (490–741) | 313 (261–358) |
| Platelet count, $\times 10^9/L$<br>(NR; 150–370) | 1605 | 218 (213–225) | 198 (186–206) | 226 (219–234) |
| IL-6, pg/mL<br>(NR, 0–3,4) | 90 | 28.4 (18.8–40.1) | 30.9 (27.2–244) | 26 (17.9–43.9) |
| Lactate dehydrogenase,<br>U/L (NR, 100–190) | 1458 | 321 (315–330) | 380 (364–408) | 308 (301–316) |
| Procalcitonin, ng/mL<br>(NR, 0–0.5) | 637 | 0.12 (0.1–0.14) | 0.4 (0.3–0.5) | 0.09 (0.08–0.11) |
| C-reactive protein, mg/L<br>(NR, 0–5) | 1482 | 71.1 (65.6–77.4) | 137.7<br>(118.1,153.8) | 57.8 (51.8–62) |
| Prothrombin time, s | 1581 | 11.1 (11.1–11.2) | 11.5 (11.4–11.7) | 10.9 (10.9–11) |
| High-sensitivity cardiac troponin I, ng/L<br>(NR, 0–53.5) | 253 | 11.2 (9.1–18.2) | 66.9 (29.7–115.2) | 8.2 (6.4–11) |
| **ICU cohort** |  | **n=75** | **n=55** | **n=20** |
| Lymphocyte count,<br>×10<sup>9</sup>/L<br>(NR 1.1–4.5) | 72 | 0.85 (0.77–0.97) | 0.77 (0.66–0.85) | 0.98 (0.87–1.15) |
| ALT, U/L<br>(NR, 0–35) | 53 | 38 (32–51) | 35 (31–52) | 47 (29–73) |
| Albumin g/dL<br>(NR, 2.9–5.2) | 28 | 4.1 (4.0–4.3) | 4.1 (4–4.4) | 4.3 (4–4.4) |
| Creatine kinase, U/L<br>(NR, 40–280) | 33 | 122 (102–176) | 122 (99–176) | 135 (107–329) |
| Creatinine mg/dL<br>(NR, 0.7–1.3) | 64 | 0.89 (0.8–1) | 0.92 (0.82–1.16) | 0.85 (0.73–0.97) |
| D-dimer, ng/mL<br>(NR, 0–500) | 26 | 1075 (752–1256) | 1088 (745–1824) | 758 (752–1647) |
| Serum ferritin, ng/L<br>(NR, 22–322) | 3 | 817 (520–852) | 835 (817–852) | 520 |
| Platelet count, $\times 10^9/L$<br>(NR, 150–370) | 65 | 185 (170–224) | 185 (165–225) | 205 (175–265) |
| IL-6, pg/mL<br>(NR, 0–3.4) | 3 | 30.9 (11.5–244.0) | 30.9 (11.5–244) | - |
| Lactate dehydrogenase,<br>U/L (NR, 100–190) | 33 | 365 (323–451) | 388 (332–520) | 328 (240–401) |
| Procalcitonin, ng/mL<br>(NR, 0–0.5) | 17 | 0.2 (0.1–0.5) | 0.38 (0.12–0.93) | 0.08 (0.06–0.16) |
| C-reactive protein, mg/L<br>(NR, 0–5) | 58 | 139.8 (109.8–188.4) | 151 (118.1–203) | 123.8 (25.1–169.7) |
| Prothrombin time, s | 63 | 11.2 (11.0–11.4) | 11.3 (11–11.5) | 11.1 (10.5–11.4) |
| High-sensitivity cardiac troponin I, ng/L<br>(NR, 0–53.5) | 7 | 24.6 (11.2–92.7) | 15.4 (11.2–73.4) | 80.2 (67.2–92.7) |

Abbreviations: ALT, alanine aminotransferase; IL-6, interleukin-6; IQR, interquartile range; NR, normal range.)

## DISCUSSION

To the best of our knowledge, this is the first report of a large cohort of patients hospitalized with COVID-19 in Europe. Our cohort includes all consecutive patients with a final outcome (discharge or death) admitted to our hospital during the heaviest COVID-19 case load on Madrid’s hospital system.

Similar to other cohorts^9-15^, hospitalized patients in Madrid were elderly and had numerous comorbidities, the most common of which were arterial hypertension and diabetes. Our male/female ratio was 0.9, which is lower than the 1.5 reported in the series from Wuhan (China)^12^ and New York City^10^. In Madrid, the male/female ratio for individuals older than 60 and 75 years was 0.74 and 0.61, respectively^16^. Differences compared with other cohorts could be partially explained by the different male/female ratio in the Madrid population pyramid. Despite our lower male/female ratio compared with other reports, the mortality for each age group was notably higher for the male patients than for the female patients, as reported in other cohorts. It is relevant that one third of the patients included in our cohort were nursing homes residents.

The overall mortality (20.7%) in our series by age group was similar to that of the New York cohort (21%)^10^ and lower than that of a Wuhan cohort (28.3%)^12^. In our cohort, older age and the presence of comorbidities were more common for the patients with fatal outcomes, both for the entire cohort and for those admitted to the ICU.

The most frequent symptoms at admission were fever, cough and dyspnea; myalgias and diarrhea were also common. Notably, 12.8% of the patients had anosmia as a presenting symptom, as described in other cohorts^17^. The time from disease onset was short (6 days), and the patients’ respiratory status on admission (as reflected by SatO_2_) was generally poor, with half of the patients presenting an SatO_2_ <95%.

The laboratory values on admission were consistent with an impaired immune-inflammatory profile characterized by lymphopenia and elevated D-dimer, procalcitonin, ferritin and C-reactive protein. Most of these abnormal laboratory readings were more common in the patients with fatal outcomes. Creatine kinase and troponin levels on admission were also higher in patients with a fatal outcome a finding also reported in other series^18^.

The majority of our patients recevied treatment with pressumed activity against SARS-CoV-2. At present there are no clinical trial data supporting that any of these treatments improve outcomes of COVIID-19

This study has a number of limitations. First, the data were collected from various databases, both manually and automatically. The data included manually in the eCRF was monitored and curated. Second, we did not conduct a follow-up of the patients after discharge. Third our reported mortality rates might change when our full cohort of hospitalized patients is analyzed

In summary, this study provides initial data on the clinical and laboratory features and outcomes of patients hospitalized with COVID-19 infection in a large teaching hospital in Madrid during the peak of the epidemic

## Data Availability

NA

## Notes

### Competing Interest Statement

The authors have declared no competing interest.

### Funding Statement

No funded

